# Associations of social media use with tobacco smoking and e-cigarettes: a national longitudinal study

**DOI:** 10.1101/2023.06.03.23290924

**Authors:** Nicholas S. Hopkinson, Charlotte Vrinten, Jennie C. Parnham, Márta K. Radó, Filippos T. Filippidis, Eszter Vamos, Anthony A. Laverty

## Abstract

**Background:** Social media may influence children and young people’s health behaviour, including smoking and e-cigarette use.

**Methods:** We analysed data from participants aged 10-25 in the UK Household Longitudinal Study 2015-2022. The amount of social media use reported on a normal weekday, was related to current tobacco smoking and e-cigarette use. Generalised Estimating Equation (GEE) logistic regression models investigated associations of social media use with tobacco and e-cigarette use. Models controlled for possible confounders including age, sex, country of UK, ethnicity, household income and use of tobacco/e-cigarettes by others within the home.

**Results:** Among 10,808 participants with 27,962 observations, current tobacco smoking was reported by 2,237(20.7%) at least one time point, and current e-cigarette use by 1,013 participants (9.4%). In adjusted GEE models, increasing use of social media was associated with greater odds of current smoking and this was particularly apparent at higher levels of use (AOR 3.11, CI 2.41-4.03 for ≥7hours/day vs no use). Associations were similar for e-cigarettes, e.g. OR=3.04, CI 2.11-4.40 for ≥7hours social media use versus none).

**Conclusions:** Social media use is associated with an increased risk of e-cigarette and tobacco use, reinforcing the need for policies to address this as an avenue for marketing to children and young people.

## INTRODUCTION

Smoking remains a major driver of inequalities in health and one of the leading preventable cause of mortality and morbidity, killing 2 in three long term users (1), although rates of tobacco smoking among adolescents and young people in the UK have been declining since the introduction of the country’s first comprehensive tobacco control plan in 1998. In recent years, the use of e-cigarettes by people trying to cut down and quit smoking has increased, with established evidence now that this is an effective quitting tool (2). However in parallel with this there has also been increasing use of e-cigarettes by children and young people, many of whom have not smoked. This raises particular concerns as vaping may cause immediate harm especially to developing lungs, likely increases the risk of long term health problems if use persists, and also may increase the risk that they go on to take up smoking. Survey data indicates differences in e-cigarette use between groups; with children in families with tobacco smokers and those in lower income households being more likely to use e-cigarettes (3)(4)(5). Understanding the mechanisms that drive e-cigarette use is key to developing strategies to prevent harm.

Substantial increases in both the proportion of young people using social media, and the time spent on them have been noted (6). Use increases with age and girls are more likely to spend longer periods of time on social media than boys (7). Social media may be driving tobacco and e-cigarette use through both direct, targeted advertisements and through the use of influencers by the tobacco industry (8). To date, most evidence on the impact of social media on tobacco and e-cigarette use has focussed on America and comprises cross-sectional studies (9)9)(10). The only two previous UK studies include a cohort study which did find that social media use at age 14 was associated with cigarette, e-cigarette, and dual use at age 17 (12). A second cross-national study from 42 countries including the UK concluded that there was a link between social media use and substance use but did not examine tobacco use separately from other substances (13). These studies however, did not have detailed assessment of potential inequality concerns, and both only contain data up to 2018.

Analyses of Instagram have identified networks of influencers promoting e-cigarettes, often without disclosing financial relationships (14). Comparative analyses in the UK have found good compliance with advertising standards for e-cigarettes on traditional media, but high levels of breaches on social media (14)(16). Any proposal to regulate social media needs to be justified and based on evidence. To contribute to this, we therefore examined the longitudinal relationship of social media use with tobacco and e-cigarette use among adolescents and young people in the UK.

## METHODS

Data come from participants of the UK Household Longitudinal Study (UKHLS), also known as Understanding Society (17). This is a longitudinal household panel study with annual household surveys that started in 2009. The original sample consisted of a clustered and stratified probability sample of approximately 28,000 households in the UK. Data are collected via face-to-face interviews carried out by a trained interviewer in the respondent’s home and via online, self-completion questionnaires. Adults over the age of 16 or above are asked to complete an individual questionnaire, including a self-completion questionnaire. Household members aged 10-15 years are asked to complete a short self-completion questionnaire, after permission from their parent or carer.

We have focused on adolescents and young adults aged 10-25 years and used data from 2015-2021 (wave 7 to wave 12). Questions on e-cigarette use were added to UKHLS in 2015. Participation in the panel is voluntary, with a gift voucher sent to encourage completion of the questionnaires and a further gift voucher sent when these are completed. All participants provided consent to be interviewed. The University of Essex Ethics Committee has approved all data collection (18).

### Outcomes and exposure

We used three separate binary outcomes: current cigarette use, current e-cigarette use and current dual use of both products. Participants were classified as current cigarette smokers if they responded: “I usually smoke between one and six cigarettes a week”, or “I usually smoke more than six cigarettes a week”. All other responses were coded as non-users. The same question was used for all waves of data and for all ages.

Current e-cigarette use was first assessed in 2009 with the question “Do you ever use electronic cigarettes (e-cigarettes)?” with response options “yes” and “no”. From wave 8 (2016/17) onwards participants were classified as current e-cigarette users if they responded: “I sometimes use e-cigarettes but less than once a month”, “I use e-cigarettes at least once a month, but less than once a week”, or “I use e-cigarettes at least once a week”. All other responses were coded as non-users.

Dual use was classified as participants currently using both products, with those using only one or no products classed as non-dual users.

The main exposure variable was social media use. Participants were first asked ‘Do you belong to any social networking web-sites?’ (yes/no), and if yes, they were also asked how many hours they spend chatting or interacting with friends through a social website on a normal weekday, with the following response options: “None”, “Less than an hour”, “1-3 hours”, “4-6 hours” and “7 or more hours”. We combined those reporting “none” along with those who were not a member of a social media website into a reference category of “not a member or no use” (7).

### Covariates

We considered a range of potentially relevant socio-demographics; age, sex, country in UK, self-defined ethnic group (collapsed into White vs. non-White due to low numbers in the non-White category), an indicator of living in an urban or rural areas (derived from Office for National Statistics Rural and Urban Classification of Output Areas), and equivalised household net income (based on OECD (Organisation for Economic Co-operation and Development) equivalence scale which was used to adjust household income by household composition (19)).

### Statistical analyses

We compared differences in socio-demographics between categories of social media use using t-tests and ANOVA. We used binary generalised estimating equation (GEE) regression models (family: binomial; link: logit; correlation matrix: exchangeable) to assess relationships between social media use and product use, developing separate models for each outcome: tobacco smoking, e-cigarette use and dual use. GEE models assess changes over time and account for the correlation caused by observations being from the same individuals (20). We also present tests for trend based on categories of social media use. Analyses adjusted for time (categorical) as well as the socio-demographic variables listed above. Models of tobacco smoking additionally adjusted for parental tobacco use, models of e-cigarette use adjusted for parental e-cigarette use, and models of dual use adjusted for both of these. Analyses used survey weights designed by the UKHLS survey team to account for the clustered and stratified probability sampling of the study and non-response bias (21).

We tested for interactions between social media use and sex, due to higher levels of social media use among women. We also tested for interactions between household income (in three groups) and social media. We present stratified analyses for both of these.

### Sensitivity analyses

We performed a range of sensitivity analyses to test the robustness of our findings. As it is possible that those not using social media at all are atypical we conducted our analyses excluding these participants. Our main analyses used household income as a marker of socio-economic status. We also performed our analyses using Index of Multiple Deprivation (in five groups) as a marker of socio-economic status. Finally, we also used fixed effects analyses to directly test if changes in social media use corresponded to uptake of tobacco smoking and e-cigarette use. These were adjusted for the time-varying variables use of tobacco/e-cigarettes by parents and household income. These models were on a smaller subset of individuals who were not product users when entering the study and who were found to change their social media use over time.

## RESULTS

Outcomes and covariates across categories of social media use are shown in **Table 1**. Overall, 8.6% of the sample reported current smoking at one or more data points, 3.8% reported current e-cigarette use, 1.7% of participants were dual users at one or more data points.

**Table 1:** Description of sample observations by social media use.

| **Weekday social media use** | **None or not a member** | **Less than an hour** | **1-3 hrs** | **4-6 hrs** | **7 or more** | **Total** | **P value for difference between groups** |
| --- | --- | --- | --- | --- | --- | --- | --- |
| Current tobacco smoking (%) | 2.0 | 6.3 | 9.2 | 12.2 | 15.7 | 8.6 | <0.001 |
| Current e-cigarette use (%) | 0.9 | 3.1 | 3.8 | 5.9 | 6.7 | 3.8 | <0.001 |
| Dual use (%) | 0.3 | 1.3 | 1.7 | 2.7 | 3.6 | 1.7 | <0.001 |
| Male (%) | 57.5 | 52.4 | 45.3 | 38.3 | 39.9 | 47.1 | <0.001 |
| Mean age (SD) | 12.0 (2.7) | 15.22 (4.0) | 16.49 (3.6) | 16.95 (3.0) | 17.7 (2.6) | 15.74 (3.8) | <0.001 |
| White ethnicity (%) | 65.6 | 71.4 | 75.1 | 75.0 | 74.2 | 72.8 | <0.001 |
| Urban areas (%) | 79.4 | 78.1 | 77.4 | 79.6 | 81.4 | 78.5 | <0.001 |
| Mean Household income (£) (SD) | 1679 (3485) | 1709 (1290) | 1689 (1055) | 1622 (1066) | 1551 (960) | 1670 (1666) | <0.001 |
| Parental tobacco use (%) | 17.0 | 16.3 | 18.6 | 21.0 | 25.2 | 18.7 | <0.001 |
| Parental e-cigarette use (%) | 7.3 | 7.5 | 8.3 | 9.4 | 10.5 | 8.3 | <0.001 |
| Overall N | 4,356 | 7,808 | 11,739 | 5,110 | 2,715 | 31,728 |  |

Tobacco smoking, e-cigarette use and dual use were all more common among participants reporting greater use of social media (all p<0.001). 2.0% of participants who did not use social media reported being a current smoker compared with 15.7% among those using social media for seven or more hours per day. E-cigarette use ranged from 0.9% among those not using social media to 6.7% among those using it for seven or more hours per day.

Differences between categories of social media use were apparent for all variables studied (all p<0.001). In particular males were less likely to be in higher social media us groups (57.5% of the ‘none or no social media group’ compared with 39.9% of the ‘7 or more hours per day’ group). Social media use increased with age (mean age of ‘none or no social media group’ 12.0 years vs. 17.7 years for the ‘7 or more hours per day’ group). Parental tobacco use was more common among those using social media more frequently (17.3% for the ‘none or no social media group’ vs. 25.2% for the ‘7 or more hours per day’ group), as was parental e-cigarette use (7.3% and 10.5% respectively).

**Table 2** shows results of our GEE models of social media use and tobacco smoking. Tobacco use was more common among those using social media more frequently (p for trend <0.001). Those using social media for ‘less than an hour a day’ were more likely to be current tobacco users than those using social media ‘none or not a member’ (Adjusted Odds Ratio = 1.92, p=<0.01) (Table 2). Those using social media for ‘7 or more hours per day’ were substantially more likely to be current tobacco users (AOR = 3.60, p<0.01).

**Table 2:**
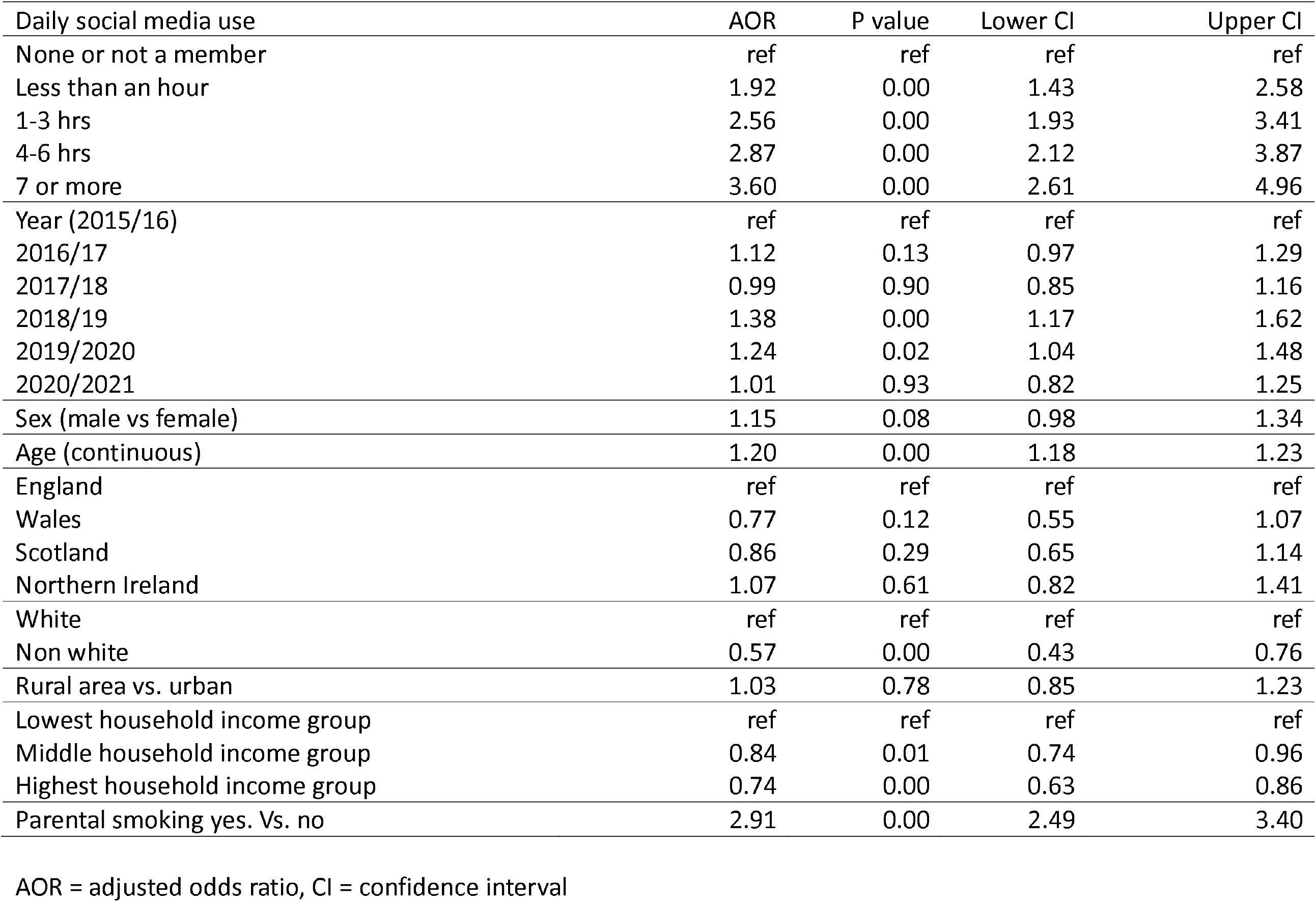
Associations of current tobacco use from Generalised Estimating Equation model

**Table 3** shows results for e-cigarette use. E-cigarette use was more common among those using social media more frequently (p for trend <0.001). E-cigarette use was more common among those using social media ‘less than an hour per day’ compared with those using it ‘none or not at all’ (AOR=1.70, 95% CI 1.09 to 2.66). E-cigarette use was considerably more likely among participants using social media ‘7 or more hours per day’ (AOR = 3.65, 95%CI 2.18 to 6.11).

**Table 3:** Associations of current e-cigarette use from Generalised Estimating Equation model

| Daily social media use | AOR | P value | Lower CI | Upper CI |
| --- | --- | --- | --- | --- |
| None or not a member | ref | ref | ref | ref |
| Less than an hour | 1.70 | 0.02 | 1.09 | 2.66 |
| 1-3 hrs | 2.16 | 0.00 | 1.35 | 3.44 |
| 4-6 hrs | 3.45 | 0.00 | 2.14 | 5.55 |
| 7 or more | 3.65 | 0.00 | 2.18 | 6.11 |
| Year (2015/16) | ref | ref | ref | ref |
| 2016/17 | 0.48 | 0.00 | 0.37 | 0.62 |
| 2017/18 | 0.54 | 0.00 | 0.43 | 0.68 |
| 2018/19 | 0.79 | 0.06 | 0.63 | 1.01 |
| 2019/2020 | 0.86 | 0.25 | 0.66 | 1.12 |
| 2020/2021 | 1.05 | 0.75 | 0.79 | 1.38 |
| Sex (male vs female) | 2.07 | 0.00 | 1.68 | 2.54 |
| Age (continuous) | 1.19 | 0.00 | 1.16 | 1.23 |
| England | ref | ref | ref | ref |
| Wales | 0.68 | 0.08 | 0.44 | 1.05 |
| Scotland | 0.72 | 0.06 | 0.51 | 1.01 |
| Northern Ireland | 0.88 | 0.50 | 0.62 | 1.27 |
| White | ref | ref | ref | ref |
| Non white | 0.60 | 0.01 | 0.41 | 0.86 |
| Rural area vs. urban | 1.05 | 0.66 | 0.83 | 1.33 |
| Lowest household income group | ref | ref | ref | ref |
| Middle household income group | 0.84 | 0.10 | 0.67 | 1.04 |
| Highest household income group | 0.78 | 0.04 | 0.63 | 0.98 |
| Parental E-cigarette use yes. Vs. no | 2.78 | 0.00 | 2.18 | 3.54 |

AOR = adjusted odds ratio, CI = confidence interval

**Table 4** shows results for dual tobacco and e-cigarette use. These models have wide confidence intervals reflecting low levels of dual use. Those using social media more frequently were more likely to be dual users (p for trend<0.001). Those using social media ‘less than an hour per day’ compared with those using it ‘none or not at all’ were more likely to be dual users (AOR=2.25, 95% CI 0.98 to 5.14). Dual use was more likely among participants using social media ‘7 or more hours per day’ (AOR = 6.23, 95%CI 2.62 to 14.86).

**Table 4:**
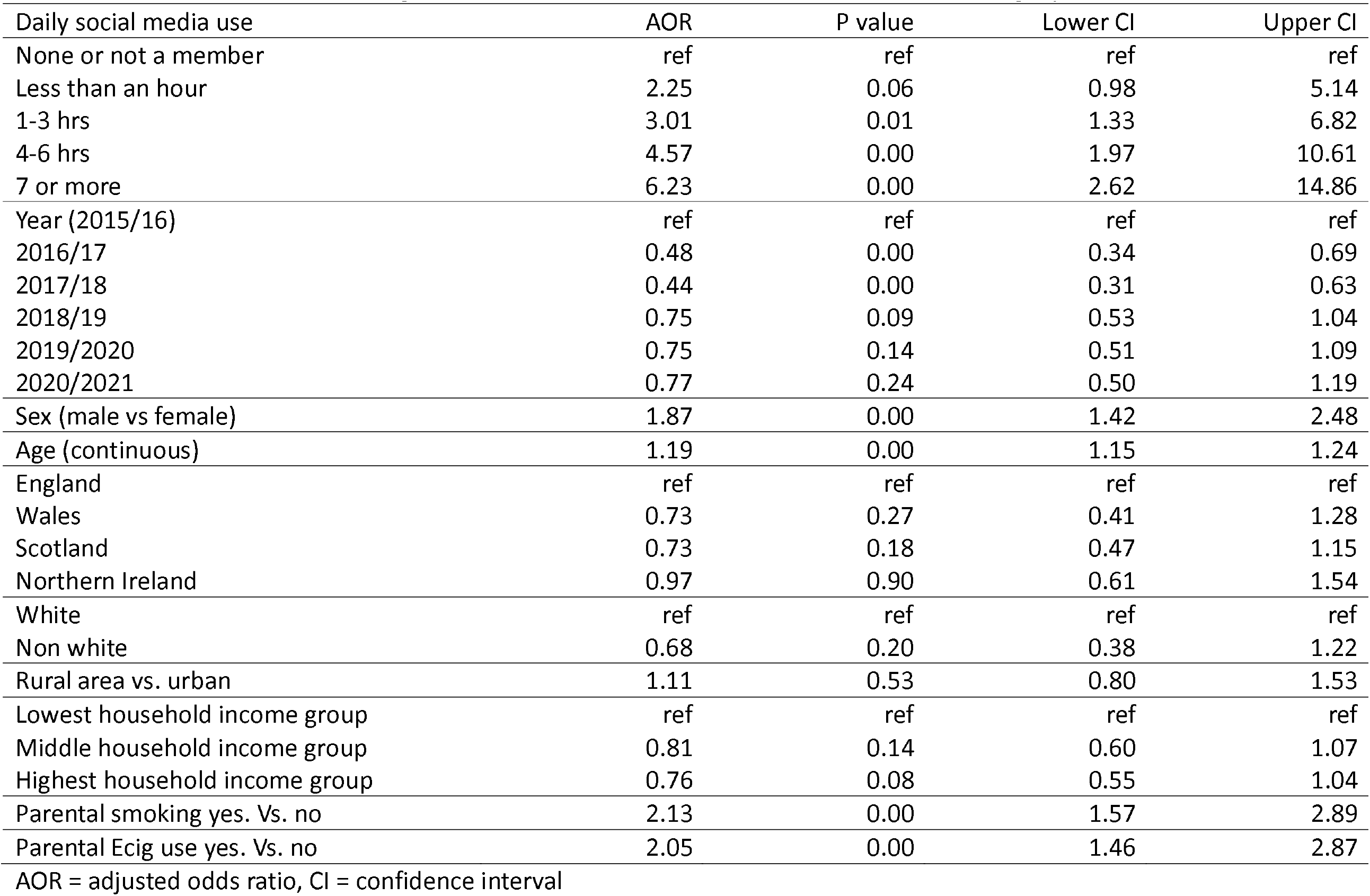
Associations of current e-cigarette and tobacco dual use from Generalised Estimating Equation model

Interactions of social media and sex were statistically significant for both tobacco and e-cigarettes (both p<0.001). In stratified models (**Table 5**) adjusted odds ratios were similar between the sexes for current tobacco use (e.g. AOR for ‘7 or more hours of social media per day’ 3.72, 95%CI 2.35 to 5.90 for males, vs. AOR 3.47, 95%CI 2.22 to 5.44 for females). For e-cigarettes, point estimates for females were higher than for males, although confidence intervals overlapped (AOR for ‘7 or more hours of social media per day’ 2.72, 95%CI 1.10 to 6.70 for males vs. AOR 4.56, 95%CI 2.46 to 8.46 for females).

**Table 5:** Associations of current e-cigarette and tobacco from gender stratified Generalised Estimating Equation models

| Daily social media use | Current tobacco use |  |  |  |  |  |
| --- | --- | --- | --- | --- | --- | --- |
|  | Males |  |  | Females |  |  |
| None or not a member | ref | ref | ref | ref | ref | ref |
| less than an hour | 2.01 | 1.30 | 3.10 | 1.83 | 1.23 | 2.74 |
| 1-3 hrs | 2.59 | 1.71 | 3.91 | 2.52 | 1.70 | 3.75 |
| 4-6 hrs | 2.92 | 1.90 | 4.49 | 2.81 | 1.85 | 4.27 |
| 7 or more | 3.72 | 2.35 | 5.90 | 3.47 | 2.22 | 5.44 |
| Daily social media use | Current e-cigarette use |  |  |  |  |  |
|  | Males |  |  | Females |  |  |
| None or not a member | ref | ref | ref | ref | ref | ref |
| less than an hour | 1.45 | 0.67 | 3.16 | 1.92 | 1.12 | 3.30 |
| 1-3 hrs | 1.50 | 0.62 | 3.62 | 2.73 | 1.61 | 4.62 |
| 4-6 hrs | 2.74 | 1.17 | 6.39 | 3.98 | 2.26 | 7.00 |
| 7 or more | 2.72 | 1.10 | 6.70 | 4.56 | 2.46 | 8.46 |

Results from models controlled for year, age, sex, country in UK, self-defined ethnic group (White vs. non-White), an indicator of living in an urban or rural areas, and equivalised household net income. Tobacco use models were adjusted for tobacco use by caregivers and e-cigarette models by use of e-cigarettes by caregivers.

Interactions with household income categories were statistically significant (p<0.001 for both tobacco and e-cigarettes) (**Table 6**). In stratified analyses of tobacco smoking, point estimates for the richest income group were higher than for the lowest income group, although confidence intervals overlapped. E.g. (AOR for ‘7 or more hours of social media per day’ 5.22, 95%CI 2.82 to 9.67 for richest income group, vs. AOR 4.17, 95%CI 2.27 to 7.65 for the lowest income group). This pattern was similar for current e-cigarette use (e.g. (AOR for ‘7 or more hours of social media per day’ 12.34, 95%CI 3.87 to 39.37 for richest income group, vs. AOR 2.61, 95%CI 1.20 to 5.72 for the lowest income group).

**Table 6:**
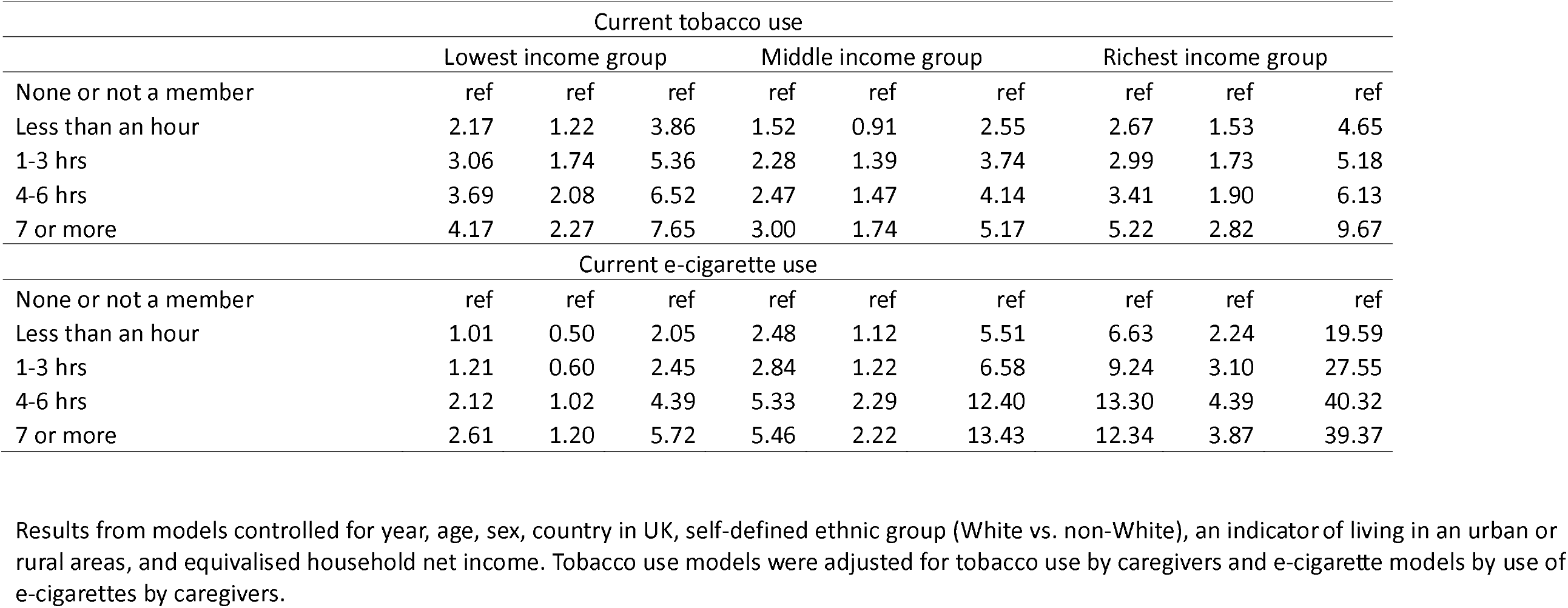
Associations of current e-cigarette and tobacco from household income stratified Generalised Estimating Equation models Current tobacco use

| Current tobacco use |  |  |  |  |  |  |  |  |  |
| --- | --- | --- | --- | --- | --- | --- | --- | --- | --- |
|  | Lowest income group |  |  | Middle income group |  |  | Richest income group |  |  |
| None or not a member | ref | ref | ref | ref | ref | ref | ref | ref | ref |
| Less than an hour | 2.17 | 1.22 | 3.86 | 1.52 | 0.91 | 2.55 | 2.67 | 1.53 | 4.65 |
| 1-3 hrs | 3.06 | 1.74 | 5.36 | 2.28 | 1.39 | 3.74 | 2.99 | 1.73 | 5.18 |
| 4-6 hrs | 3.69 | 2.08 | 6.52 | 2.47 | 1.47 | 4.14 | 3.41 | 1.90 | 6.13 |
| 7 or more | 4.17 | 2.27 | 7.65 | 3.00 | 1.74 | 5.17 | 5.22 | 2.82 | 9.67 |
| Current e-cigarette use |  |  |  |  |  |  |  |  |  |
| None or not a member | ref | ref | ref | ref | ref | ref | ref | ref | ref |
| Less than an hour | 1.01 | 0.50 | 2.05 | 2.48 | 1.12 | 5.51 | 6.63 | 2.24 | 19.59 |
| 1-3 hrs | 1.21 | 0.60 | 2.45 | 2.84 | 1.22 | 6.58 | 9.24 | 3.10 | 27.55 |
| 4-6 hrs | 2.12 | 1.02 | 4.39 | 5.33 | 2.29 | 12.40 | 13.30 | 4.39 | 40.32 |
| 7 or more | 2.61 | 1.20 | 5.72 | 5.46 | 2.22 | 13.43 | 12.34 | 3.87 | 39.37 |

Results from models controlled for year, age, sex, country in UK, self-defined ethnic group (White vs. non-White), an indicator of living in an urban or rural areas, and equivalised household net income. Tobacco use models were adjusted for tobacco use by caregivers and e-cigarette models by use of e-cigarettes by caregivers.

### Sensitivity analyses

GEE analyses excluding those not using any social media were similar to our main analyses (**Appendix Table 1**). Analyses using IMD as a marker of socio-economic status rather than household income also gave similar results (**Appendix Table 2**).

Fixed effect analyses gave similar results to main analyses for uptake of tobacco smoking (**Appendix Table 3**). It should be noted that sample size was much reduced for this model (n=864). These found some evidence that changes in social media use are linked to uptake of tobacco smoking in a dose-response manner (p for trend=0.053). For example, changing to using social media for 7 or more hours per day was associated with more than five times the odds of taking up tobacco smoking (AOR = 5.67, p<0.01).

Associations between changes in social media use and uptake of e-cigarettes did not reveal associations between changes in social media use and uptake of e-cigarettes. These had even lower sample sizes (n=564). For example, adjusted odds ratios of e-cigarette uptake ranged from 0.86 (95%CI 0.46 to 1.62) for participants using social media ‘less than an hour a day’ to AOR=1.24, 95%CI 0.64 to 2.40, for those using social media ‘7 or more hours per day.’ The test for trend in odds ratios was not significant (p=0.423).

## DISCUSSION

The main finding of the present study is that in children and young adults, increasing social media use was associated with a higher likelihood of both current smoking and current use of e-cigarettes. This association was independent of other factors associated with increasing smoking and vaping including age, gender, socioeconomic status and parental smoking and vaping.

There are a number of possible, non-exclusive explanations for this relationship. First, and most straightforwardly, there is evidence that the corporations behind smoking and vaping make use of social media to advertise and promote their products. This includes direct advertising which is algorithmically targeted and the use of paid social media influencers who present smoking and vaping as a fashionable and desirable activity. Greater time spent on social media is likely to increase exposure to this form of influence, increasing the extent to which product use is perceived as a norm. Second, social media use has been shown to have features in common with reward-seeking addictive behaviour (22). High social media use may increase susceptibility to other addictive behaviours like smoking. Alternatively, both behaviours may be driven by a common susceptibility. Thirdly, as a space that is largely unsupervised by parents/caregivers social media use may encourage behaviours that are transgressive, including smoking and vaping. There is evidence that peer smoking is a strong influence on child uptake of smoking (12)(23) and social media is one of the ways in which peer-smoking and vaping will be experienced both by seeing others behaviour and by sharing “influencer content” that promotes these behaviours.

### Limitations

The data that we collected relates only to the quantity of social media use. We do not have information about which platforms were being used or how individuals were using them. For example, the extent to which they are interacting socially with individuals they know or consuming content from influencers, personalities or media corporations etc. As well as quantifying this there is a need for more in depth qualitative investigation into how young people are experiencing social media content in relation to smoking and vaping.

### Policy implications

The content that social media users are exposed to is to a substantial extent algorithmically controlled, both through targeted advertising and by the promotion of material that maximises engagement in order to increase revenue to the platform. This can be controlled. For example, far right imagery which is otherwise widely available is largely inaccessible in Germany, as a consequence of German law which social media platforms are bound to enforce. As such companies have substantial power to modify exposure to material that promotes smoking and vaping if they choose to or are compelled to. Voluntary codes seem unlikely to achieve this, and the introduction and enforcement on bans on material that promote this should be considered.

Policies to reduce overall social media use by young people are beyond the scope of this paper and it is certainly the case that for many people social media use helps to build networks and is beneficial. Nevertheless, for parent and policy makers, measures to reduce the very high usage levels described by some participants in this study may represent an area for interventions to improve health and well-being in children and young people.

## Data Availability

Data from this study are from the UK Household Longitudinal Study and available from the UK Data Service https://ukdataservice.ac.uk

## Appendix

**Appendix Table 1:** GEE models excluding participants not using social media (main effects only)

| Weekday social media use | AOR | Lower CI | Upper CI |
| --- | --- | --- | --- |
| Current tobacco smoking |  |  |  |
| less than an hour | ref | ref | ref |
| 1-3 hrs | 1.33 | 1.15 | 1.53 |
| 4-6 hrs | 1.47 | 1.23 | 1.76 |
| 7 or more | 1.86 | 1.52 | 2.27 |
| Current e-cigarette use |  |  |  |
| less than an hour | ref | ref | ref |
| 1-3 hrs | 1.28 | 1.01 | 1.63 |
| 4-6 hrs | 2.07 | 1.58 | 2.71 |
| 7 or more | 2.20 | 1.59 | 3.04 |
| Current dual use (tobacco and e-cigarettes) |  |  |  |
| less than an hour | ref | ref | ref |
| 1-3 hrs | 1.36 | 0.96 | 1.93 |
| 4-6 hrs | 2.06 | 1.39 | 3.06 |
| 7 or more | 2.84 | 1.84 | 4.40 |

**Appendix Table 2:** GEE models using IMD as marker of socio-economic status

| Weekday social media use | AOR | Lower CI | Upper CI |
| --- | --- | --- | --- |
| Current tobacco smoking |  |  |  |
| None or not a member | ref | ref | ref |
| less than an hour | 1.94 | 1.45 | 2.61 |
| 1-3 hrs | 2.62 | 1.97 | 3.49 |
| 4-6 hrs | 2.94 | 2.18 | 3.96 |
| 7 or more | 3.65 | 2.65 | 5.02 |
| Current e-cigarette use |  |  |  |
| None or not a member | ref | ref | ref |
| less than an hour | 1.72 | 1.10 | 2.69 |
| 1-3 hrs | 2.18 | 1.37 | 3.47 |
| 4-6 hrs | 3.54 | 2.20 | 5.68 |
| 7 or more | 3.64 | 2.18 | 6.08 |
| Current dual use (tobacco and e-cigarettes) |  |  |  |
| None or not a member | ref | ref | ref |
| less than an hour | 2.34 | 1.04 | 5.28 |
| 1-3 hrs | 3.15 | 1.41 | 7.04 |
| 4-6 hrs | 4.76 | 2.08 | 10.88 |
| 7 or more | 6.56 | 2.80 | 15.37 |

**Appendix Table 3:** Fixed effect models of changes in social media use and uptake of tobacco smoking and e-cigarettes Note these from two separate models controlled for changes in household income (continuous) and in tobacco/e-cigarette use by parents. In tobacco model N = 864, in e-cigarette model N = 564.

| Current tobacco use |  |  |  |  |
| --- | --- | --- | --- | --- |
| None or not a member | ref | ref | ref | ref |
| less than an hour | 1.69 | 0.07 | 0.96 | 2.99 |
| 1-3 hrs | 2.10 | 0.01 | 1.21 | 3.66 |
| 4-6 hrs | 2.01 | 0.02 | 1.13 | 3.56 |
| 7 or more | 2.33 | 0.01 | 1.28 | 4.24 |
| Current e-cigarette use |  |  |  |  |
| None or not a member | ref | ref | ref | ref |
| less than an hour | 0.86 | 0.65 | 0.46 | 1.62 |
| 1-3 hrs | 0.97 | 0.92 | 0.53 | 1.79 |
| 4-6 hrs | 1.15 | 0.66 | 0.61 | 2.16 |
| 7 or more | 1.24 | 0.52 | 0.64 | 2.40 |

